# Help-Seeking Behavior and Its Associated Factors Among 18– 24 years Victims of Intimate Partner Violence in Kenya: Insights from a Respondent-Driven Survey

**DOI:** 10.1101/2024.11.27.24318074

**Authors:** Humwa Felix, Onguru Daniel, Memiah Peter, Asito S. Amolo

## Abstract

**Background:** Intimate partner violence (IPV) is a pervasive issue with significant mental health implications. Understanding the factors that influence help-seeking behavior among IPV survivors is crucial for developing effective interventions.

**Objective:** This study aims to identify the demographic, educational, behavioral, mental health, and social network characteristics associated with help-seeking behavior among IPV survivors aged between 18-24 years.

**Methods:** This was a cross-sectional respondent-driven survey conducted within three cities of Kisumu, Mombasa, and Nairobi in Kenya among individuals aged 18-24 years. Data was analyzed using univariate and multivariable logistic regression to identify factors associated with help-seeking behavior.

**Results:** Data was analyzed from 351 (58% females) participants who were exposed to IPV. Residency, educational attainment, mental health status, social networks, and health status significantly impact the likelihood of seeking help (p < .05).

**Conclusion:** The study highlights the complex interplay of various factors influencing help-seeking behavior among IPV survivors. These findings underscore the need for targeted interventions that address specific barriers and facilitators of help-seeking behavior, particularly in urban settings.

## Introduction

Intimate partner violence (IPV) is a significant public health issue that affects millions of individuals globally. It encompasses physical, sexual, and emotional abuse by a current or former partner, leading to severe mental and physical health consequences for survivors [1]. The prevalence of IPV is alarmingly high, and has been reported to be at 27% globally [2]. Sub Saharan Africa is impacted by IPV and studies report that its prevalence ranges between 10% and 60% [3, 4, 5]. The impact of IPV extends beyond immediate physical injuries, contributing to long-term mental health issues such as depression, anxiety, post-traumatic stress disorder (PTSD), and suicidal ideation [2, 6].

Help-seeking behavior among IPV survivors is influenced by a complex interplay of factors, including demographic characteristics, educational attainment, mental health status, and social networks [7]. Demographic factors such as age, gender, and location significantly impact an individual’s likelihood of seeking help. For instance, urban residents may have better access to mental health resources and face less stigma associated with seeking help compared to their rural counterparts [8]. Additionally, educational attainment plays a critical role, with higher levels of education often associated with increased awareness and utilization of mental health services [7].

Mental health status is a crucial determinant of help-seeking behavior among IPV survivors. Individuals experiencing severe mental health issues, such as suicidal thoughts or a history of abuse, may be more likely to seek help due to the higher level of distress and need for support [2]. However, stigma and fear of judgment can act as significant barriers, preventing many survivors from disclosing their experiences and seeking the necessary help [9]. Creating supportive environments that encourage disclosure and reduce stigma is essential for improving help-seeking behavior among IPV survivors [9].

Social networks and relationships also play a pivotal role in help-seeking behavior. Strong social support networks can facilitate help-seeking by providing emotional support and practical assistance [10]. Conversely, individuals with limited or negative social networks may be less likely to seek help due to a lack of support and increased feelings of isolation. The rise of social media has added a new dimension to social networks, with online relationships potentially influencing help-seeking behavior in both positive and negative ways [10].

Understanding the complex interplay of factors influencing help-seeking behavior among IPV survivors is essential for developing effective interventions and support systems. This study aims to identify the demographic, educational, behavioral, mental health, and social network characteristics associated with help-seeking behavior among IPV survivors. By addressing the specific barriers and facilitators of help-seeking behavior, we can improve access to mental health resources and support for IPV survivors, ultimately enhancing their overall well-being and quality of life [7, 11, 12].

## Methods

### Aim

The aim of this study was to examine help-seeking behavior and its associated factors among respondents aged 18-24 years who experienced IPV in Kenya.

### Setting

The research study used data collected in three cities, Nairobi Mombasa and Kisumu, in the Republic of Kenya. Nairobi, the capital city, spans approximately 696 square kilometers, Mombasa and Kisumu covers about 294.7 417 square kilometers respectively [13]. According to the Kenya National Bureau of Statistics (KNBS), the population of individuals aged 18-24 years in these cities is significant, with Nairobi having a substantial youth demographic. Economically, Nairobi is a hub for various sectors including technology, finance, and manufacturing, with a notable presence in the digital and gig economy. Mombasa’s economy is heavily influenced by its strategic position as a port city, focusing on shipping, tourism, and the blue economy Kisumu, on the other hand, leverages its proximity to Lake Victoria, with significant activities in fishing, agriculture, and trade [14].

The youth in these cities face several health-related challenges that include mental health issues such as depression and anxiety are prevalent, particularly among out-of-school adolescents [15]. Additionally, challenges like early pregnancies, unsafe abortions, and gender-based violence are common [16, 17]. These health issues highlight the need for targeted interventions to support the well-being of young people in these urban areas.

### Sample

The data utilizes collected data from the REACH (Reaching, Engaging Adolescents and Young Adults for Care Continuum in Health) respondent-driven survey. The interactive mobile app can be downloaded from Google Play or the Apple Store and could be used offline only to communicate with the REACH web application once the survey was completed. The survey combines existing screening tools, adapted for use jointly with AYA in Kenya through a co-design process and had 10 structured modules [17]. The data was collected among females and males aged between 15-26 years in the cities of Nairobi, Mombasa and Kisumu Kenya (n=1,467).

For this research, males and females aged 18-24 years who reported as ever had sex, responded to the question to have experienced intimate partner violence were included in the study (n=744). From this sample, only respondents who responded either a “Yes” or “Not comfortable disclosing” on the ever-experiencing IPV variable were selected for the research analysis (n = 351).

### Measurements

The study variables categories of interests included: 1) Help-seeking after experiencing IPV, 2) social network and friendship ties, 3) mental health characteristics 4) socio-demographic and clinical characteristics.

The dependent variable, help-seeking, was derived from a variable that asked the respondents on whether they sought help after experiencing IPV. Respondents who responded to *“I reported it”* were considered to have sought help and the other categories of *“Too embarrassed to talk to anyone about it”, “Didn’t know who to tell”* and *“Just let it go”* were considered to not to have sought help.

The social network construct variables included on whether they consider their social media friends to be close (*No/A few/Majority/All of them)*, groups of people they mostly hang out with (M*ixed crowd/Opposite sex/Same sex*) and if they get along with teachers in school (*None/One /Some/Most/All*). The friendship-ties construct was a “*yes/no”* composite variables from questions that asked whether their friends criticize them, how often they let them down and if they often the get the respondents angry.

The mental health construct consisted of questions that asked how many of the respondents become violent (*Never/Sometimes/Most of the time/Always*) and ever had suicidal thoughts (Y*es/No/Not comfortable disclosing*). Behavioral variables included questions on whether they are currently sexually active, condom use and participation in sports activities. The socio-demographic characteristics included variables such as sex, age, education, occupation while the clinical characteristics were variables on ever had an STI, HIV status, under any medication and whether they consider themselves healthy in overall.

### Statistical analysis

Data was analyzed using R version 4.2, employing descriptive statistics, Wilcoxon rank-sum tests, and Pearson chi-square tests with Rao-Scott’s correction. The study used data of individuals who were exposed to IPV. The study outcome was defined as those who sought any formal or informal help after experiencing IPV. Bivariate and multivariable logistic regression models were used to determine factors associated with the help seeking behavior among those who were exposed to IPV. Variables that had a p <0.1 in the bivariate analysis were fit in the multivariable logistic regression model. Statistical significance was taken at p < 0.05 for all analyses.

### Ethics statement

Approval for this study was obtained from both the Ethics and Scientific Review Committee of Amref Health Africa in Kenya and the Institutional Review Board of the University of West Florida. All ethical procedures were conducted and maintained throughout the study period. Participants provided an informed consent before participating in the study.

## Results

A total of 351 participants (58% female) aged 18-24 years participated in the study. The prevalence of help-seeking behavior was found to be 78 (22%).

### Help-seeking by participant characteristics

Table 1 shows the distribution of help seeking behavior by demographic, clinical, behavioral, social network and friendship perceptions. The distribution of sex was equal, with 58% female and 42% male in both groups. The median age was 22 years across all groups, and the age categories showed no significant differences. Regarding living with both parents, 57% of participants were not living with both parents, with a slightly higher percentage among those who sought help (69%) compared to those who did not (53%). A higher percentage of those who considered themselves health, did not seek help (65%) compared to those who sought help (49%) and almost similar results were observed among those who considered their guardians/siblings healthy overall. Respondents who were on medication had higher rates of seeking help (18%) compared to those who were not. A higher percentage of those who did not seek help had never thought about hurting or killing themselves (45%) compared to those who sought help (32%). Furthermore, 56% of those who sought help were not comfortable disclosing abuse history, compared to 36% of those who did not seek help. The study observed that participants whose friends criticized them, often let them done and often made them angry had higher rates of not seeking help. Additionally, in terms of social media relationships, those who sought help were less likely to consider their social media friends as close friends (30%) compared to those who did not seek help (45%). (Table 1).

**Table 1:**
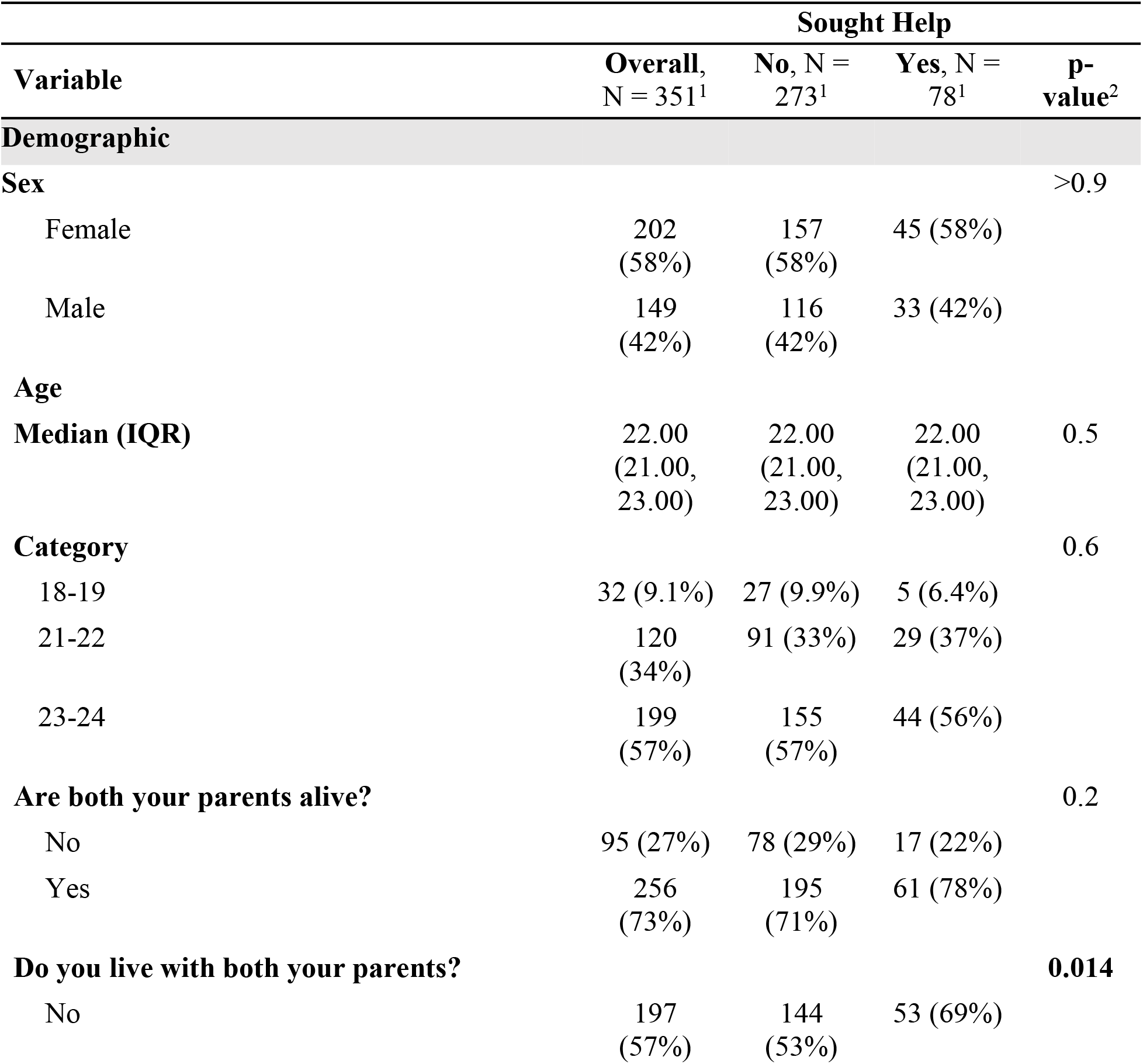
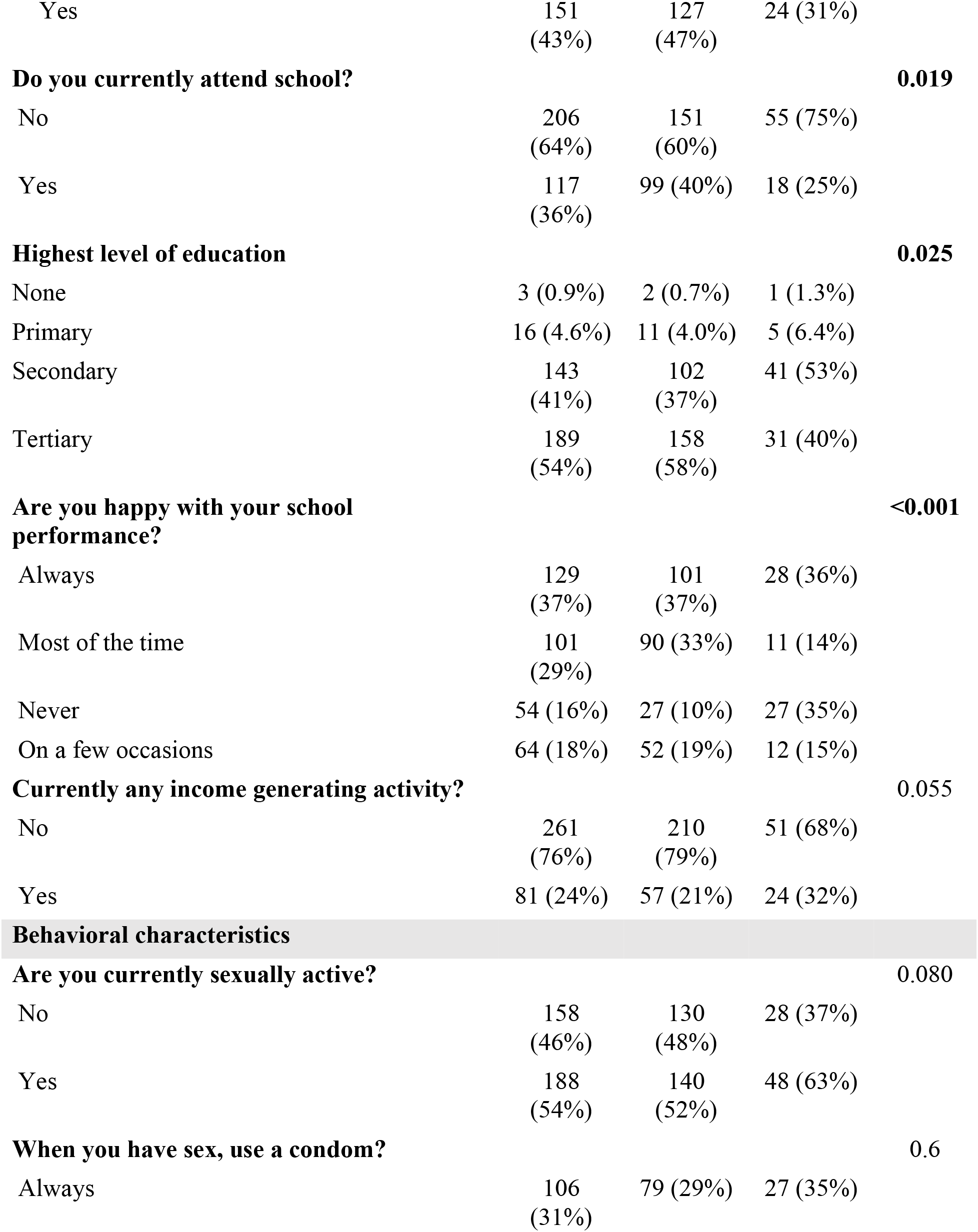
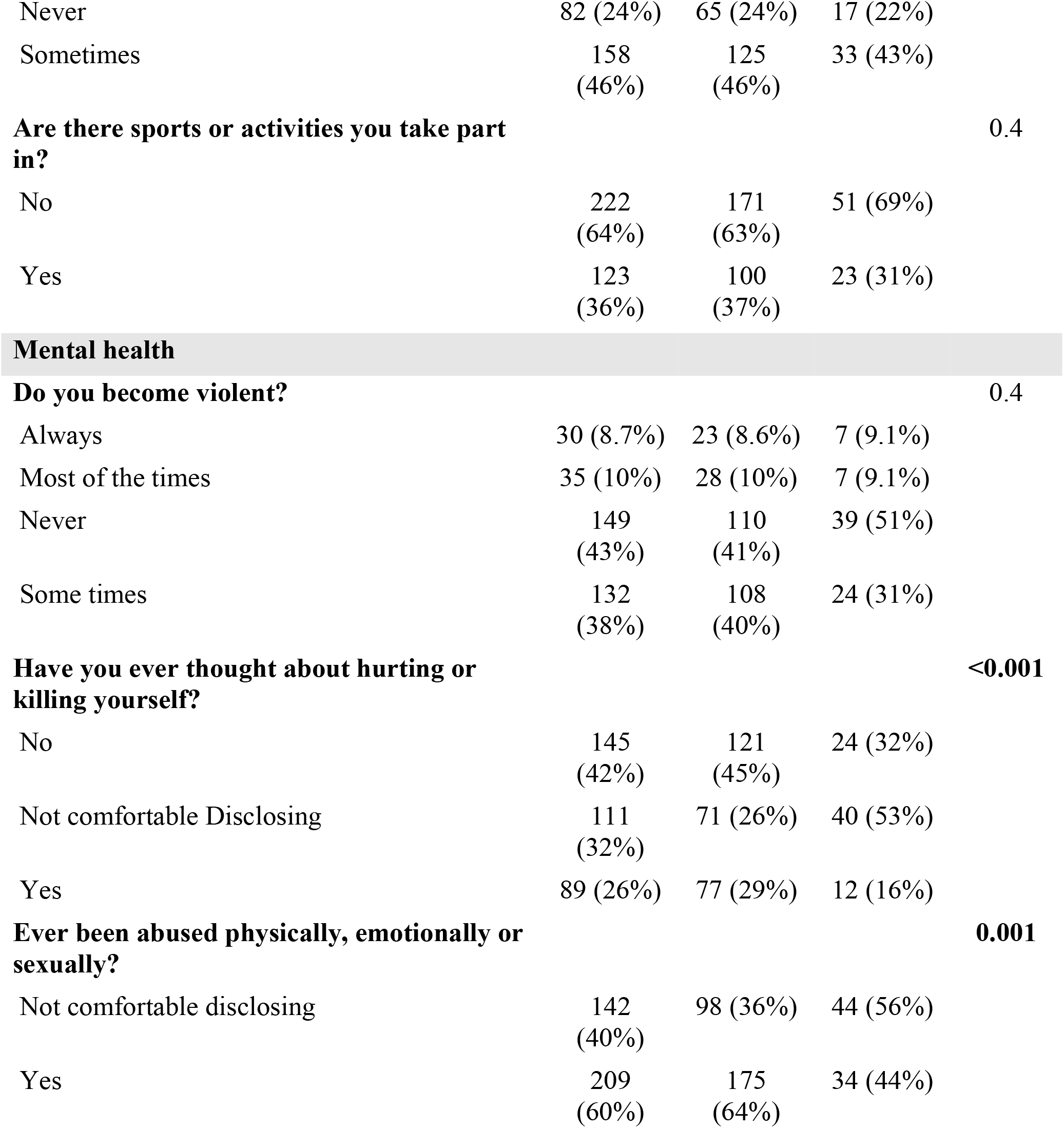
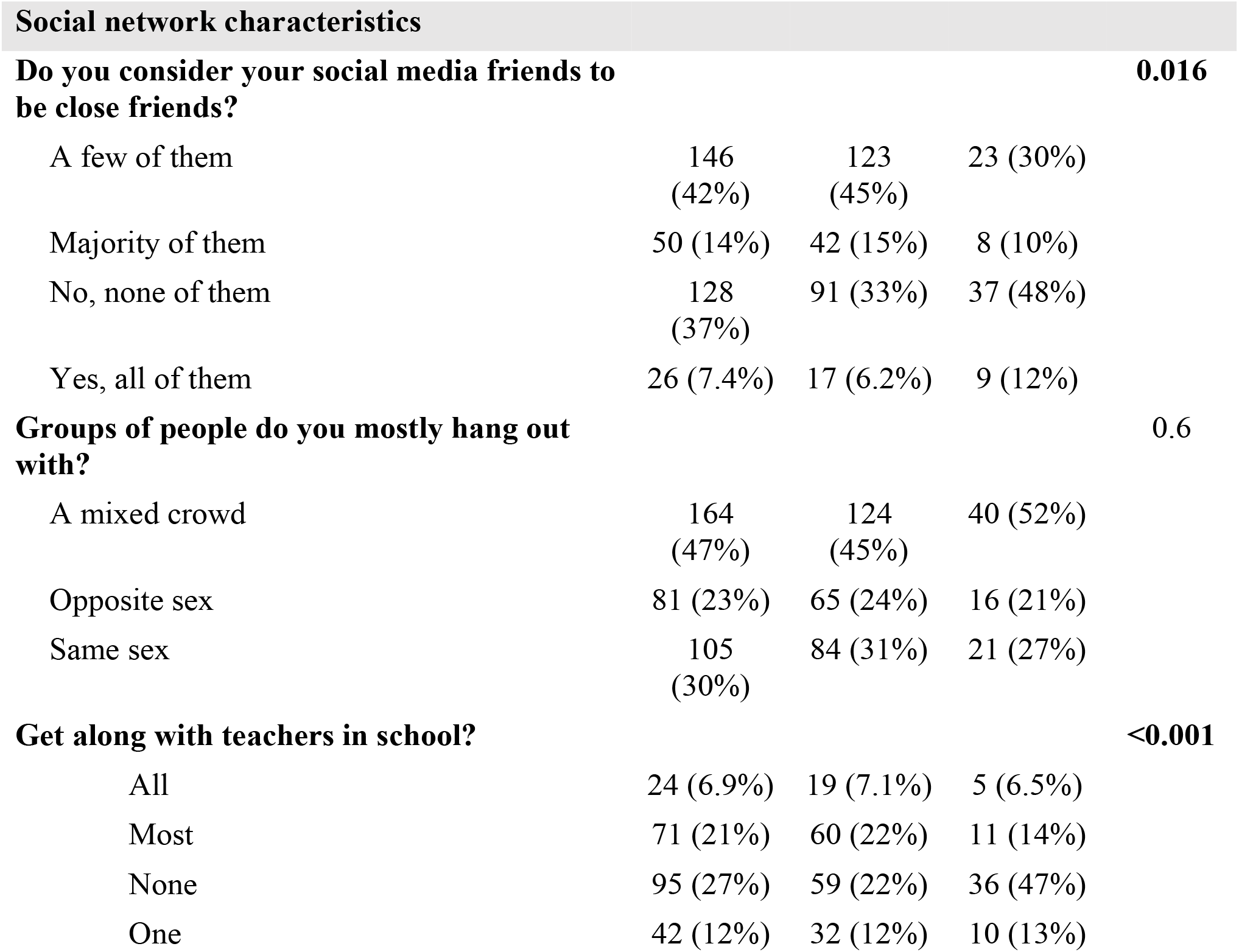
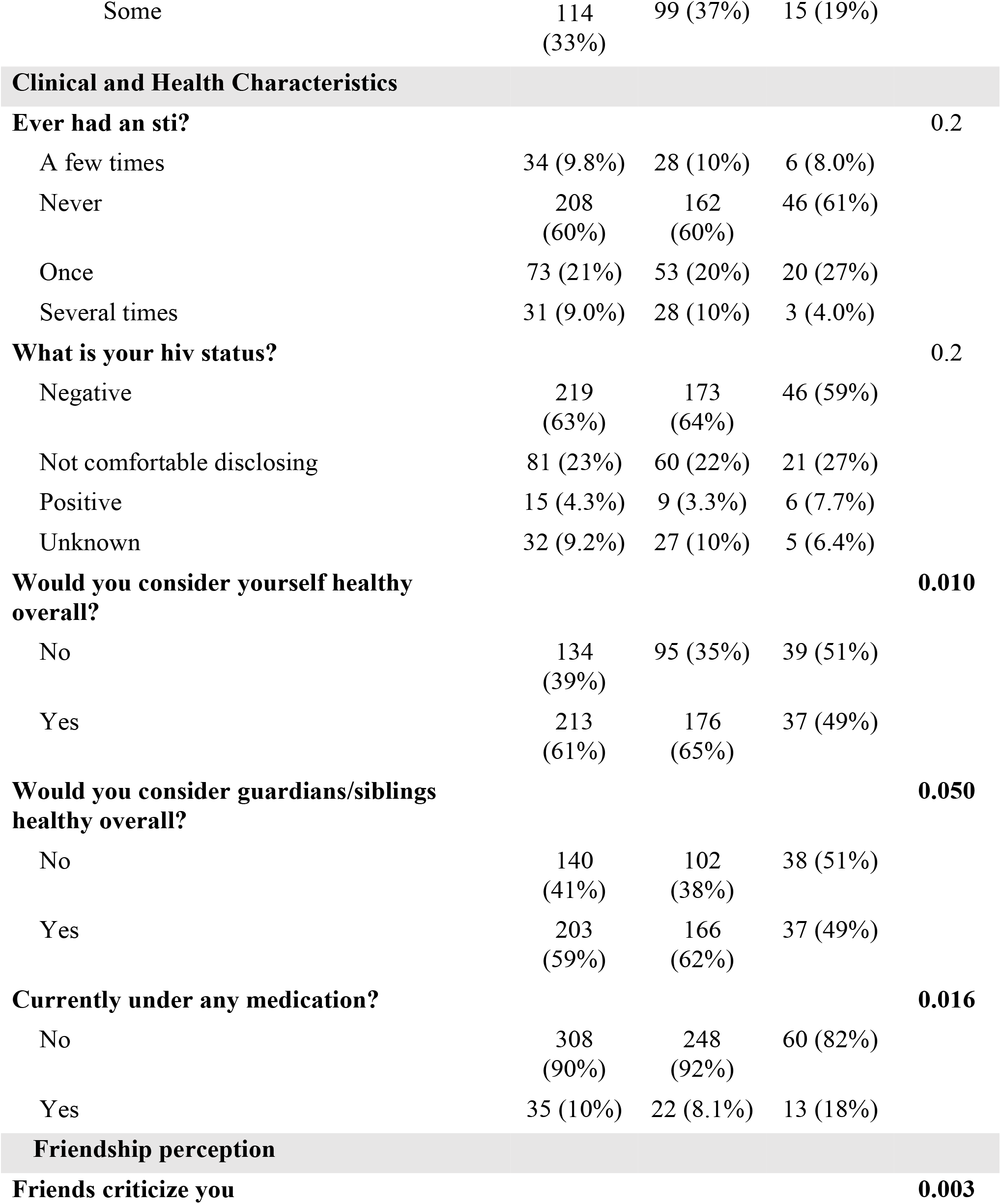
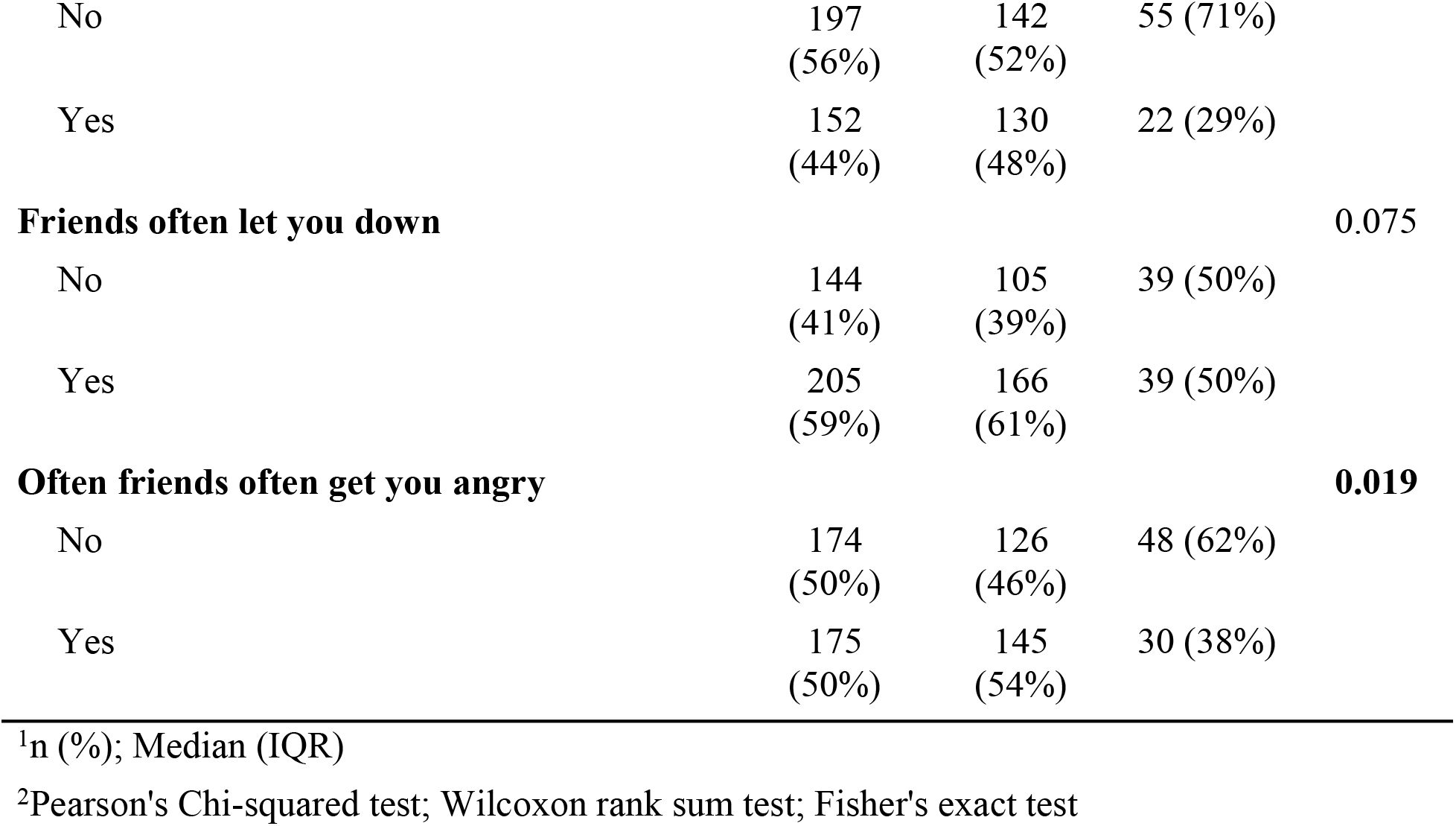
Characteristics of study participants who experienced intimate partner violence.

### Bivariate analysis

Individuals currently residing in Nairobi (OR = 4.30, 95% CI: 1.74, 13.0, *P* < .001) and Mombasa (OR = 1.52, 95% CI: 0.58, 4.75, *P* < .001) had significantly higher odds of the outcome compared to those in Kisumu. Having higher education levels were found to be associated with less help-seeking behavior as those with tertiary education exhibited lower odds compared to individuals with no education (OR = 0.39, 95% CI: 0.04, 8.60, *P* = 0.04). We also observed that individuals who were never happy with their school performance had significantly higher odds (OR = 3.61, 95% CI: 1.84, 7.17, *P* < .001) compared to those who were always happy. Conversely, those who were happy most of the time had lower odds (OR = 0.44, 95% CI: 0.20, 0.91, *P* < .001). Furthermore, individuals not comfortable disclosing thoughts of self-harm had higher odds (OR = 2.84, 95% CI: 1.59, 5.15, *P* < .001) compared to those who had never considered it. Other significant findings included the lower odds among individuals who had been abused (OR = 0.43, 95% CI: 0.26, 0.72, *P* = .001) and those who considered themselves healthy (OR = 0.51, 95% CI: 0.31, 0.86, *P* = .011). Additionally, individuals currently under medication had higher odds (OR = 2.44, 95% CI: 1.14, 5.07, *P* = .02). More results are as shown in table 2.

**Table 2:**
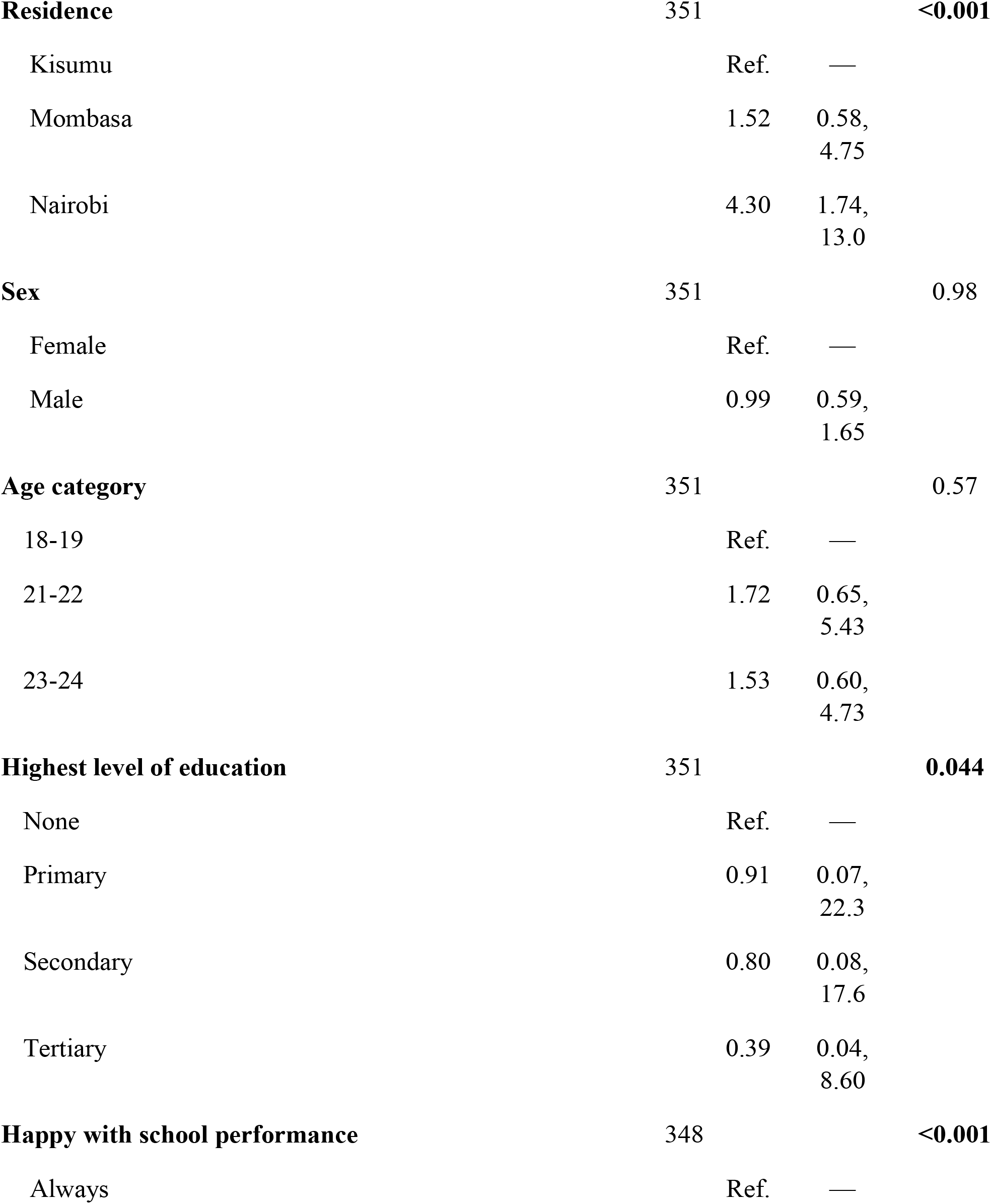
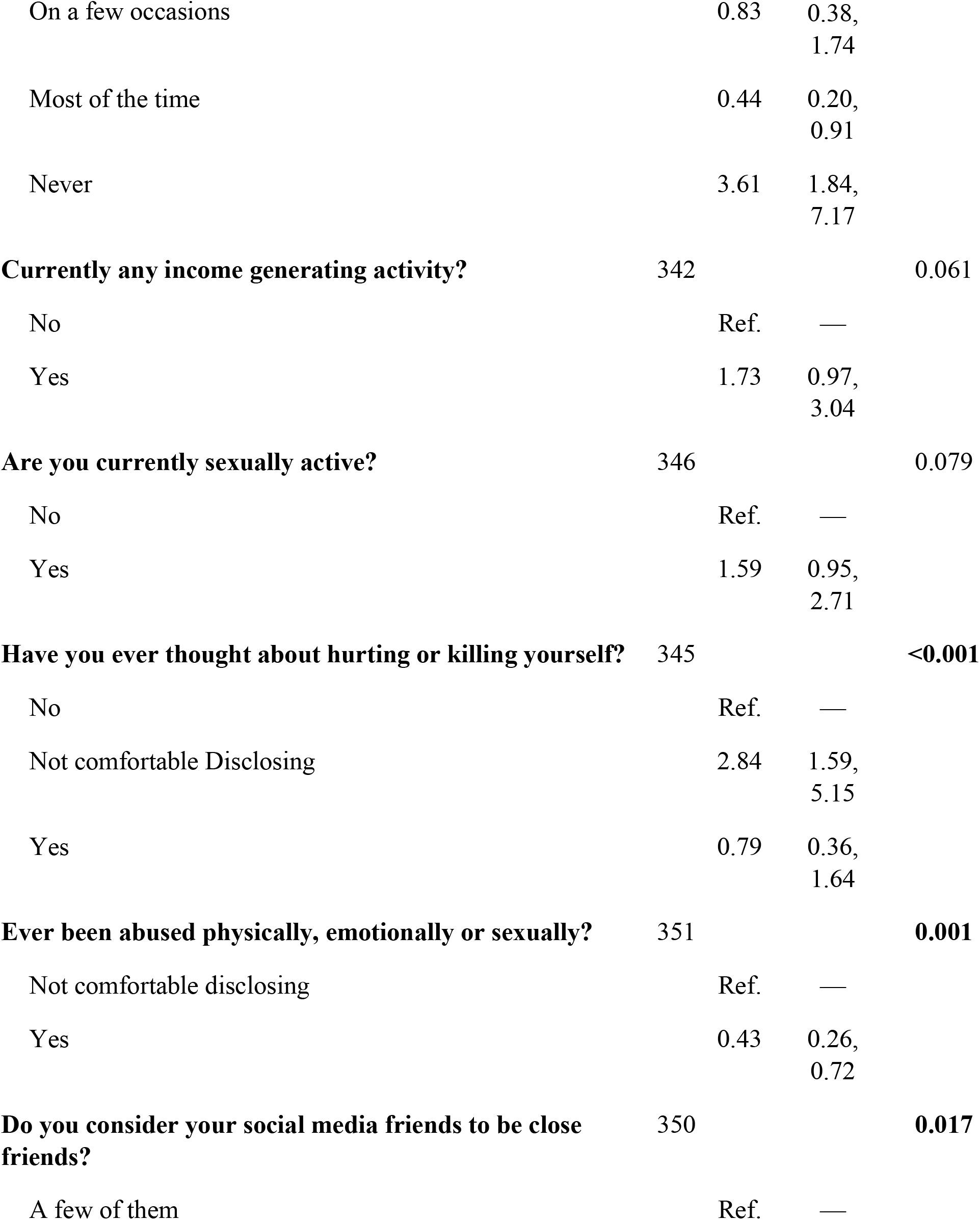
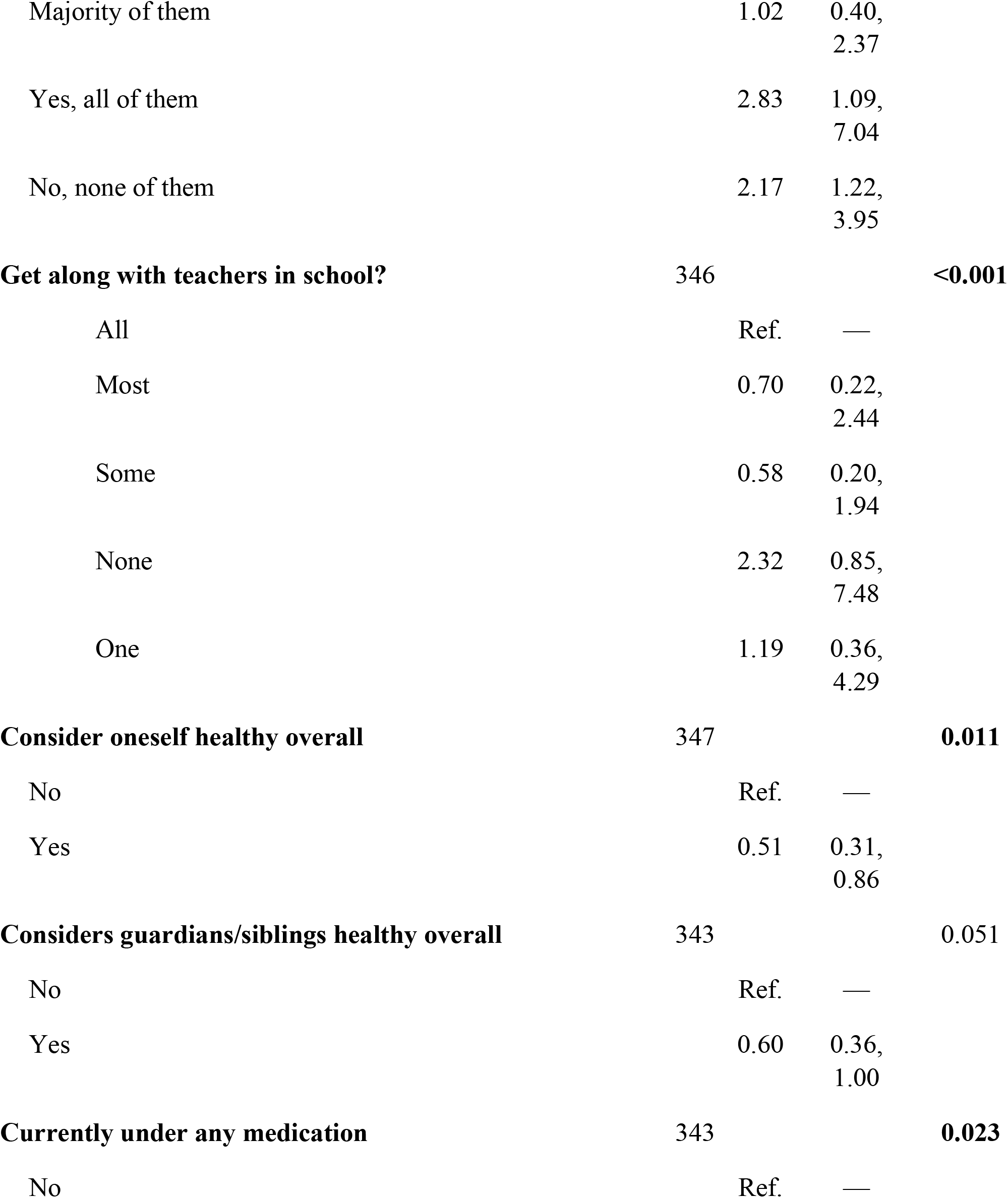
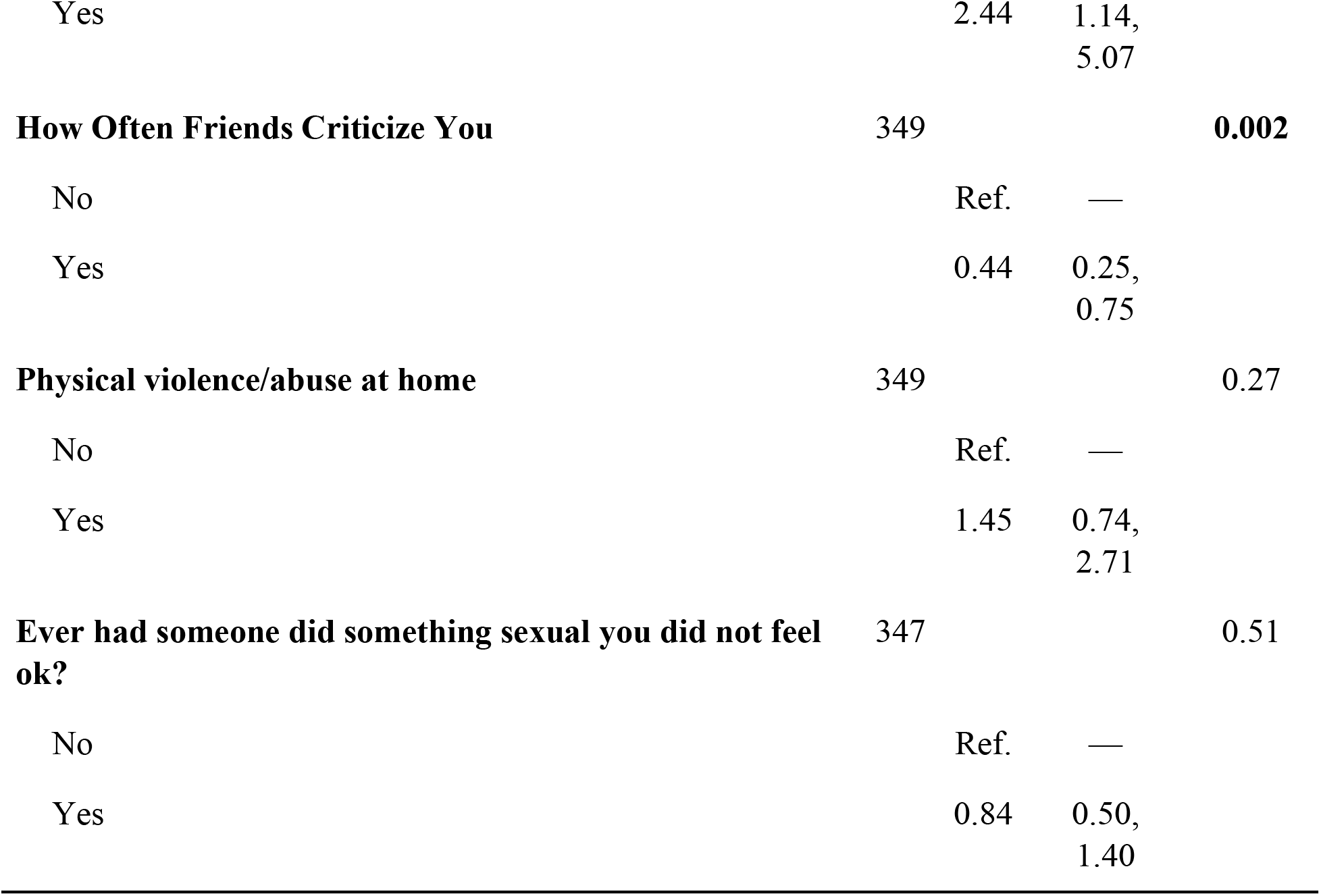
Bivariate logistic regression on factors associated with help-seeking behavior among 18–24-year-olds who experienced IPV.

| Characteristic | Univariate |  |  |  |
| --- | --- | --- | --- | --- |
|  | N | OR | 95% CI | P-value |
| **Residence** | 351 |  |  | **<0.001** |
| Kisumu |  | Ref. | — |  |
| Mombasa |  | 1.52 | 0.58, 4.75 |  |
| Nairobi |  | 4.30 | 1.74, 13.0 |  |
| **Sex** | 351 |  |  | 0.98 |
| Female |  | Ref. | — |  |
| Male |  | 0.99 | 0.59, 1.65 |  |
| **Age category** | 351 |  |  | 0.57 |
| 18-19 |  | Ref. | — |  |
| 21-22 |  | 1.72 | 0.65, 5.43 |  |
| 23-24 |  | 1.53 | 0.60, 4.73 |  |
| **Highest level of education** | 351 |  |  | **0.044** |
| None |  | Ref. | — |  |
| Primary |  | 0.91 | 0.07, 22.3 |  |
| Secondary |  | 0.80 | 0.08, 17.6 |  |
| Tertiary |  | 0.39 | 0.04, 8.60 |  |
| **Happy with school performance** | 348 |  |  | **<0.001** |
| Always |  | Ref. | — |  |

| Characteristic | N | Univariate |  |  |
| --- | --- | --- | --- | --- |
|  |  | OR | 95% CI | P-value |
| On a few occasions |  | 0.83 | 0.38,<br>1.74 |  |
| Most of the time |  | 0.44 | 0.20,<br>0.91 |  |
| Never |  | 3.61 | 1.84,<br>7.17 |  |
| **Currently any income generating activity?** | 342 |  |  | 0.061 |
| No |  | Ref. | — |  |
| Yes |  | 1.73 | 0.97,<br>3.04 |  |
| **Are you currently sexually active?** | 346 |  |  | 0.079 |
| No |  | Ref. | — |  |
| Yes |  | 1.59 | 0.95,<br>2.71 |  |
| **Have you ever thought about hurting or killing yourself?** | 345 |  |  | <0.001 |
| No |  | Ref. | — |  |
| Not comfortable Disclosing |  | 2.84 | 1.59,<br>5.15 |  |
| Yes |  | 0.79 | 0.36,<br>1.64 |  |
| **Ever been abused physically, emotionally or sexually?** | 351 |  |  | 0.001 |
| Not comfortable disclosing |  | Ref. | — |  |
| Yes |  | 0.43 | 0.26,<br>0.72 |  |
| **Do you consider your social media friends to be close friends?** | 350 |  |  | 0.017 |
| A few of them |  | Ref. | — |  |
|  | N | OR | 95% CI | P-value |
| Majority of them |  | 1.02 | 0.40,<br>2.37 |  |
| Yes, all of them |  | 2.83 | 1.09,<br>7.04 |  |
| No, none of them |  | 2.17 | 1.22,<br>3.95 |  |
| **Get along with teachers in school?** | **346** |  |  | **<0.001** |
| All |  | Ref. | — |  |
| Most |  | 0.70 | 0.22,<br>2.44 |  |
| Some |  | 0.58 | 0.20,<br>1.94 |  |
| None |  | 2.32 | 0.85,<br>7.48 |  |
| One |  | 1.19 | 0.36,<br>4.29 |  |
| **Consider oneself healthy overall** | **347** |  |  | **0.011** |
| No |  | Ref. | — |  |
| Yes |  | 0.51 | 0.31,<br>0.86 |  |
| **Considers guardians/siblings healthy overall** | **343** |  |  | **0.051** |
| No |  | Ref. | — |  |
| Yes |  | 0.60 | 0.36,<br>1.00 |  |
| **Currently under any medication** | **343** |  |  | **0.023** |
| No |  | Ref. | — |  |

| Characteristic | N | Univariate |  |  |
| --- | --- | --- | --- | --- |
|  |  | OR | 95% CI | P-value |
| Yes |  | 2.44 | 1.14, 5.07 |  |
| **How Often Friends Criticize You** | 349 |  |  | **0.002** |
| No |  | Ref. | — |  |
| Yes |  | 0.44 | 0.25, 0.75 |  |
| **Physical violence/abuse at home** | 349 |  |  | 0.27 |
| No |  | Ref. | — |  |
| Yes |  | 1.45 | 0.74, 2.71 |  |
| **Ever had someone did something sexual you did not feel ok?** | 347 |  |  | 0.51 |
| No |  | Ref. | — |  |
| Yes |  | 0.84 | 0.50, 1.40 |  |

### Multivariable analysis

Those residing in Nairobi were significantly more likely to seek help compared to those in Kisumu, with an adjusted odds ratio (aOR) of 5.24 (95% CI: 1.40, 24.6; *P* = .02). Participants who reported that they get along with some of their teachers were less likely to seek help compared to those who get along with all of their teachers (aOR = 0.20, 95% CI: 0.05, 0.84; *P* = .02). Mental health characteristics significantly influenced help-seeking behavior. Participants who reported experiencing physical violence or abuse at home were more likely to seek help (aOR = 3.08, 95% CI: 1.06, 5.11; *P* = .04). Social network characteristics also played a role; participants who considered all their social media friends to be close friends were significantly more likely to seek help (aOR = 9.12, 95% CI: 2.30, 37.3; *P* = .002). However, participants whose friends often criticized them were less likely to seek help (aOR = 0.41, 95% CI: 0.17, 0.92; *P* = .04) compared to those who were not criticized. Self-perceived health status and medication use were significant predictors of help-seeking behavior. Participants who were currently under medication were more likely to seek help (aOR = 4.09, 95% CI: 1.47, 6.7; *P* = .01). (Figure 1).

**Figure 1:**
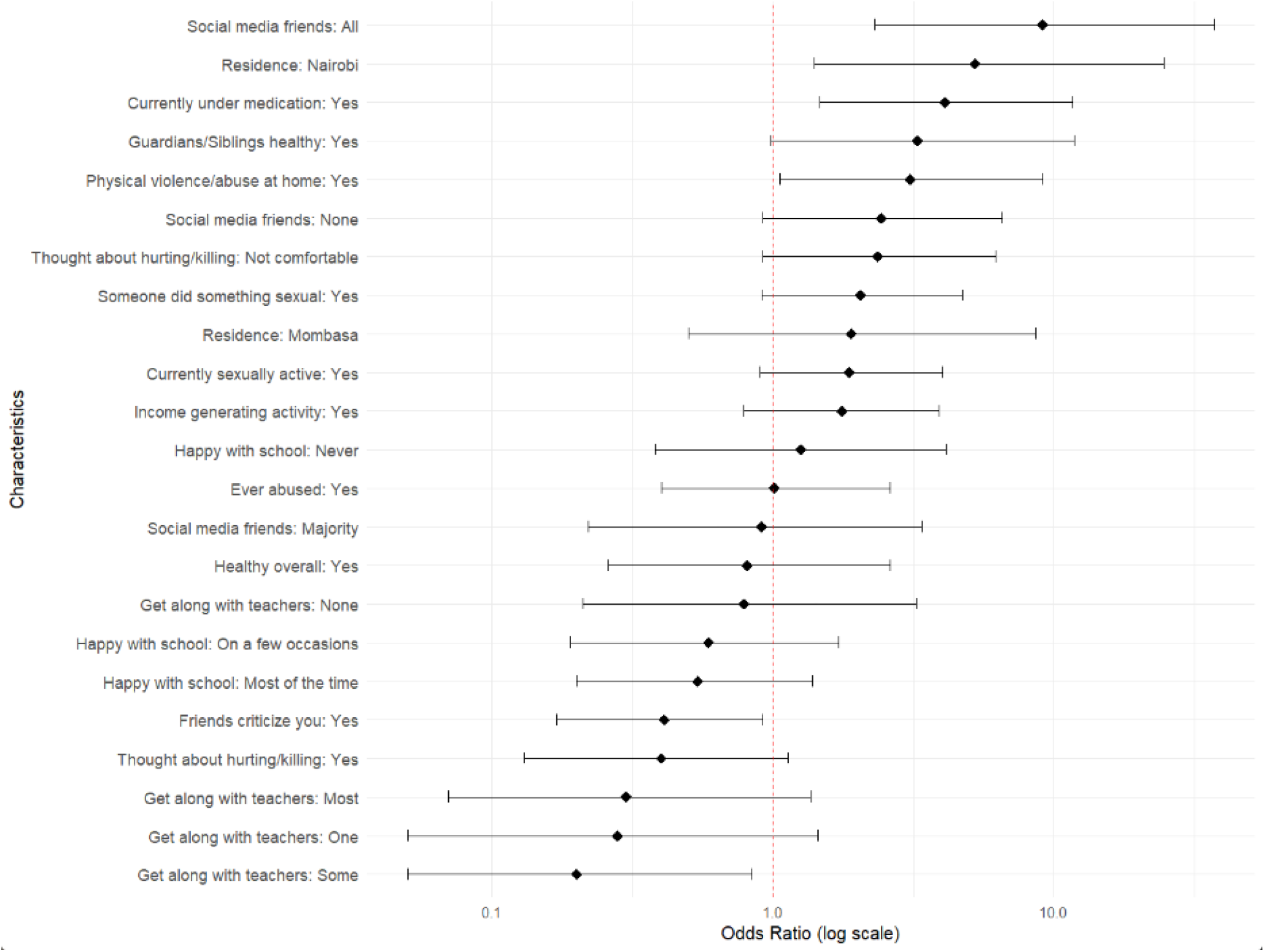
Forest plot of adjusted Odds Ratios with 95% confidence intervals.

## Discussion

The results indicate significant differences in educational, behavioral, and mental health characteristics between those who sought help and those who did not. The lack of significant gender differences in help-seeking behavior aligns with some studies but contradicts others that have found females to be more likely to seek help [18, 19]. This discrepancy may be due to the indifference in IPV among young-adults.

Educational attainment was found to be a significant factor, with participants having tertiary education being less likely to seek help. One the effects of IPV is having lower levels of education [17, 20], however this finding contrasts with some literature suggesting that higher education levels are associated with increased help-seeking behavior [21]. This indicates that the pressures and expectations associated with higher education may lead to a reluctance to seek help due to perceived stigma or fear of judgment [22].

The disparity in happiness with school performance highlights the potential impact of academic stress on mental health. Those who sought help were more likely to report never being happy with their school performance, which could be a contributing factor to their decision to seek help. Studies have shown that exposure to IPV have effects of academic underperformance because of the physical and mental health problems caused by it and it has also been linked with causing education and training disruptions [23, 24].

The mental health characteristics, particularly the higher incidence of suicidal thoughts and discomfort in disclosing abuse history among those who sought help, underscore the importance of addressing mental health stigma and providing supportive environments for disclosure. Suicidal thoughts have been found to be significantly associated with intimate partner violence [25]. These findings are supported by research that emphasizes the need for mental health interventions that are sensitive to the stigma and privacy concerns of young individuals [25, 26, 27]. The study highlights the complex interplay between educational engagement, mental health, and help-seeking behavior. Future interventions should focus on creating supportive educational environments and reducing stigma around mental health to encourage help-seeking behavior.

The role of social networks in help-seeking behavior was also evident, with participants who considered all their social media friends to be close friends being significantly more likely to seek help. This finding aligns with research suggesting that strong social support networks can facilitate help-seeking behavior [28, 29]. However, it also raises questions about the quality of social media relationships and their impact on mental health [30].

Health characteristics, including self-perceived overall health and current medication use, were significant predictors of help-seeking behavior. Participants who considered themselves healthy were less likely to seek help, which may reflect a lower perceived need for mental health services [31].Conversely, those currently under medication were more likely to seek help, indicating a higher level of engagement with healthcare services and possibly more severe mental health issues

## Conclusion

The findings suggest that understanding the contextual factors influencing young IPV victims’ help-seeking may provide useful insights for developing interventions that can mitigate the circumstances that reinforce the normalization of violence and make it difficult for them to seek help [32]They also highlighted a critical gap in the current intervention, in that the involvement of formal systems in Kenya is low, if not non-existent [33]. Given the complexity of IPV, especially in this context, there is a need for multi-strategy interventions. The first step, however, is to review the training systems given to healthcare providers, parents, and couples, and set up community systems to deal with IPV [34]. Additionally, providing a link to counseling and social workers will allow for victims’ emotional expression and support [35]

Based on these findings, the following recommendations for services can be considered: There is a need to review the training for healthcare providers and communities on IPV. Due to the unique needs among young people experiencing IPV, specific training aimed at addressing this gap may be useful. Healthcare providers should consider IPV as an underlying cause of ill health. Annual prevalence estimates for IPV across the lifespan are recommended. This should be trended so that the country is able to assess if interventions are working. Stigma surrounding the help-seeking processes and services for IPV should be addressed, and a communication strategy on services should be developed. A responsive and timely service system, depending on the presenting problem, is necessary. Young girls and boys should have access to psychosocial support services in all its spheres, depending on what they present with [36].

### Limitations

Although our findings shed light on important areas that need to be addressed, it should be noted that our study design has some limitations. This study utilized a cross-sectional sample that may limit the likelihood of generalizing findings to the general population of the adolescents and young adults (AYA). Further research into a direct relationship between help-seeking and the identified needs of young people is recommended. Longitudinal studies on help-seeking are recommended, as this is beyond the present study’s scope and design. In addition, the app was only accessible to people who could download it on their mobile phones or other technological apparatus. Thus, information from AYA who are not technologically savvy or from those who experienced technological difficulties was not collected. We should note that, despite the limitations, our findings echo those in similar settings and add to the present knowledge base on the topic. Likewise, such design has been used in the past to tap into the attitudinal survey given the stigma surrounding IPV. There is a need for a more comprehensive view on this issue. More mixed methods research is required.

## Acknowledgement and Funding

We acknowledge the university of Maryland Global Centre for Engagement (Presidential Global Impact Fund (PGIMF) which birthed this collaborative work. We are grateful to all the participants in this study. In addition, we thank the HIV Intervention Science Training Program (HISTP). This NIMH-funded multidisciplinary training program form Columbia school of social work that seeks to develop and facilitate the growth of scientists from under-represented groups conduction HIV-related dissemination and implementation research to support Prof. Memiah Peter in this study.

## Data availability statement

The datasets presented in this study can be availed on request.

## Author contributions

H.F. formulated the research question, provided general oversight of this work, authored the first draft of the manuscript, conducted data analysis, interpreted results. A.S.A., and O.D., interpreted results, reviewed several versions of the manuscripts. M.P., contributed to study design, assisted in the interpretation of results, and provided general oversight of this work. All authors reviewed this manuscript, provided feedback, and approved of the manuscript in its final form.

